# Prevalence and Age-Sex Distribution of Thyroid-stimulating Hormone Abnormalities Among Hospital Outpatients in Jashore District, Bangladesh: A Cross-Sectional Study

**DOI:** 10.64898/2026.08.16.26360524

**Authors:** Moumita Rahman Sazza, Sabrin Bashar, Md Deen Islam, Saiful Islam Noman

**Author notes:** Corresponding author, address: ECHA, Department of Pediatrics, University of Alberta, Edmonton, AB, Canada, T6G 2R3.

## Abstract

**Background:** Thyroid disorders represent a substantial endocrine disease burden across South Asia, yet systematic epidemiological data from specific regions of Bangladesh remain scarce. The objective of this study was to determine the prevalence and demographic distribution of thyroid stimulating hormone (TSH) abnormalities among hospital outpatients in Jashore district, southeastern Bangladesh.

**Methods:** A cross-sectional survey was conducted among 200 consecutive outpatients presenting for thyroid evaluation at LabAid Hospital, Jashore, from October 2017 to December 2017. Serum TSH concentrations were measured, and participants were classified as euthyroid, hypothyroid, or hyperthyroid based on standard reference intervals, then stratified by sex and seven age categories.

**Results:** Of the 200 participants (170 female, 30 male), 101 (50.5%) showed abnormal TSH values. Hypothyroidism was identified in 97 participants (48.5%) and hyperthyroidism in 4 (2.0%). The overall prevalence of thyroid dysfunction was nearly identical between sexes (female 50.6% vs male 50.0%, P=1.00), while age group showed a highly significant association with TSH abnormality (P<0.001), peaking in the 40-49-year group (89.7%). The female-to-male ratio for total thyroid dysfunction was 5.7:1, reflecting the much larger proportion of female participants rather than a higher within-sex risk.

**Conclusion:** Thyroid dysfunction, principally hypothyroidism, is highly prevalent among hospital outpatients in Jashore City, with prevalence rising sharply with age and peaking in middle adulthood, supporting the case for systematic age-targeted thyroid screening in this population.

## Background

Thyroid disorders are among the most prevalent endocrine conditions worldwide, imposing a significant burden on health systems in both developed and developing nations (1, 2). Serum thyroid stimulating hormone (TSH) measurement remains the most sensitive first-line biochemical indicator of thyroid dysfunction: elevated TSH is diagnostic of primary hypothyroidism, while suppressed TSH indicates hyperthyroidism (3, 4). Global estimates suggest that approximately 200 million individuals live with thyroid disease, with hypothyroidism being the dominant manifestation in most populations (2, 5). Females are four to ten times more susceptible to thyroid disorders than males, largely attributable to the immunomodulatory effects of estrogen on thyroid autoimmunity (6). Prevalence increases progressively with advancing age in both sexes (7, 8).

In Bangladesh, thyroid disorders represent a growing public health concern; up to 20–40% of the population may experience thyroid dysfunction over their lifetime (9). Historically, iodine deficiency was the predominant driver of hypothyroidism and endemic goiter in this region; however, the epidemiological profile has shifted toward autoimmune thyroid disease following iodine supplementation programs (10). Despite this recognized disease burden, population-level data from specific geographic regions of Bangladesh, particularly the southwestern zone, remain limited (11).

Jashore district, located in southwestern Bangladesh, represents a geographically and demographically distinct setting with potentially unique environmental and nutritional determinants of thyroid health. Understanding the local prevalence and distribution of TSH abnormalities is essential for informing context-specific screening guidelines and resource allocation. This study was therefore conducted to determine the prevalence of TSH abnormalities and to characterize their distribution by sex and age group among outpatients in Jashore, Bangladesh.

## Methods

### Study site and population

LabAid Hospital, Jashore Branch (lat 23.168, long 89.210), situated in Jashore district, Bangladesh, was selected as the study site, serving as a principal referral center for outpatient clinical laboratory services in the region. Inclusion criteria were outpatients presenting for thyroid function evaluation who provided written informed consent and a venous blood sample. Individuals currently receiving thyroid hormone replacement therapy or antithyroid medications were excluded to avoid confounding of TSH measurement.

Participants were enrolled by consecutive sampling from October to December 2017. A total of 200 participants were enrolled over the study period. Sociodemographic data, including age and sex, were recorded using a standardized, interviewer-administered questionnaire that also captured weight and location. Age was categorized into seven predefined groups: 0-19, 20-29, 30-39, 40-49, 50-59, 60-69, and ≥70 years. A 5 mL venous blood sample was collected from each participant under standardized aseptic conditions following an overnight fast of at least 8 hours.

### Detection of TSH abnormalities

The laboratory testing was performed at the LabAid Hospital clinical laboratory, Jashore, Bangladesh. Serum was separated by centrifugation at 3,000 rpm for 10 minutes and analyzed promptly. Serum TSH concentrations were determined using immunoassay methodology. Thyroid status was classified as: (i) euthyroid (TSH 0.4-4.0 mIU/L), (ii) primary hypothyroidism (TSH >4.0 mIU/L), or (iii) hyperthyroidism (TSH <0.4 mIU/L), consistent with reference intervals endorsed by the American Thyroid Association (13).

### Statistical Analysis

Data were entered and managed in Microsoft Excel (Microsoft Corporation, Office 365) and categorized according to qualitative and quantitative variables. Descriptive analysis was performed using frequencies and proportions. Tests of association between TSH status and sex or age group were performed using the chi-square test. A P-value of <0.05 was considered statistically significant.

## Results

During the three months of surveillance from October to December 2017, 200 outpatients with varied age groups and sex were screened for TSH abnormalities at LabAid Hospital, Jashore (Table 1). Of the study population, 50.5% (101/200) showed an abnormal TSH value; hypothyroidism was identified in 48.5% (97/200) and hyperthyroidism in 2.0% (4/200), while the remaining 49.5% (99/200) were classified as euthyroid (**Figure 2**).

**Table 1:** Demographic characteristics of study participants with TSH diagnosis results.

| Characteristics | Total N=200 | TSH Abnormal<br>n=101 (50.5%) | Euthyroid n=99<br>(49.5%) | P-value* |
| --- | --- | --- | --- | --- |
| **Sex** |  |  |  | 1.00 |
| Male | 30 | 15 (50.00) | 15 (50.00) |  |
| Female | 170 | 86 (50.59) | 84 (49.41) |  |
| **Age groups (years)** |  |  |  | <0.001 |
| 0-19 | 21 | 7 (33.33) | 14 (66.67) |  |
| 20-29 | 62 | 18 (29.03) | 44 (70.97) |  |
| 30-39 | 47 | 26 (55.32) | 21 (44.68) |  |
| 40-49 | 29 | 26 (89.66) | 3 (10.34) |  |
| 50-59 | 24 | 18 (75.00) | 6 (25.00) |  |
| 60-69 | 14 | 5 (35.71) | 9 (64.29) |  |
| ≥70 | 3 | 1 (33.33) | 2 (66.67) |  |
| **Total** | 200 | 101 (50.50) | 99 (49.50) |  |

\*P-value generated by chi-square test comparing TSH-abnormal vs. euthyroid counts across sex and age-group categories. $P < 0.05$ was considered statistically significant.

We examined whether sex had any effect on the prevalence of thyroid dysfunction. The cohort comprised predominantly female participants (85%, 170/200) (**Figure 1**). The prevalence of any thyroid dysfunction was nearly identical between the sexes (female 50.6% [86/170] vs male 50.0% [15/30], P=1.00, Table 1), indicating that although females carried the greater absolute case burden accounting for 83 of 97 hypothyroid cases and 3 of 4 hyperthyroid cases (female-to-male ratio 5.7:1 for total abnormal TSH, Table 2). This reflected their larger representation in the sample rather than a higher within-sex risk.

**Table 2:** Sex-stratified distribution of thyroid status among study participants (N=200)

| Thyroid Status | Male n=30 (%) | Female n=170 (%) | F:M Ratio |
| --- | --- | --- | --- |
| Hypothyroidism | 14 (7.00) | 83 (41.50) | 5.9:1 |
| Hyperthyroidism | 1 (0.50) | 3 (1.50) | 3.0:1 |
| Total abnormal TSH | 15 (7.50) | 86 (43.00) | 5.7:1 |
| Euthyroid (normal) | 15 (7.50) | 84 (42.00) | 5.6:1 |
| **Total** | 30 (15.00) | 170 (85.00) | 5.7:1 |

F:M, female-to-male ratio calculated from absolute case counts. Percentages represent the proportion of the total study population (N=200).

**Figure 1.**
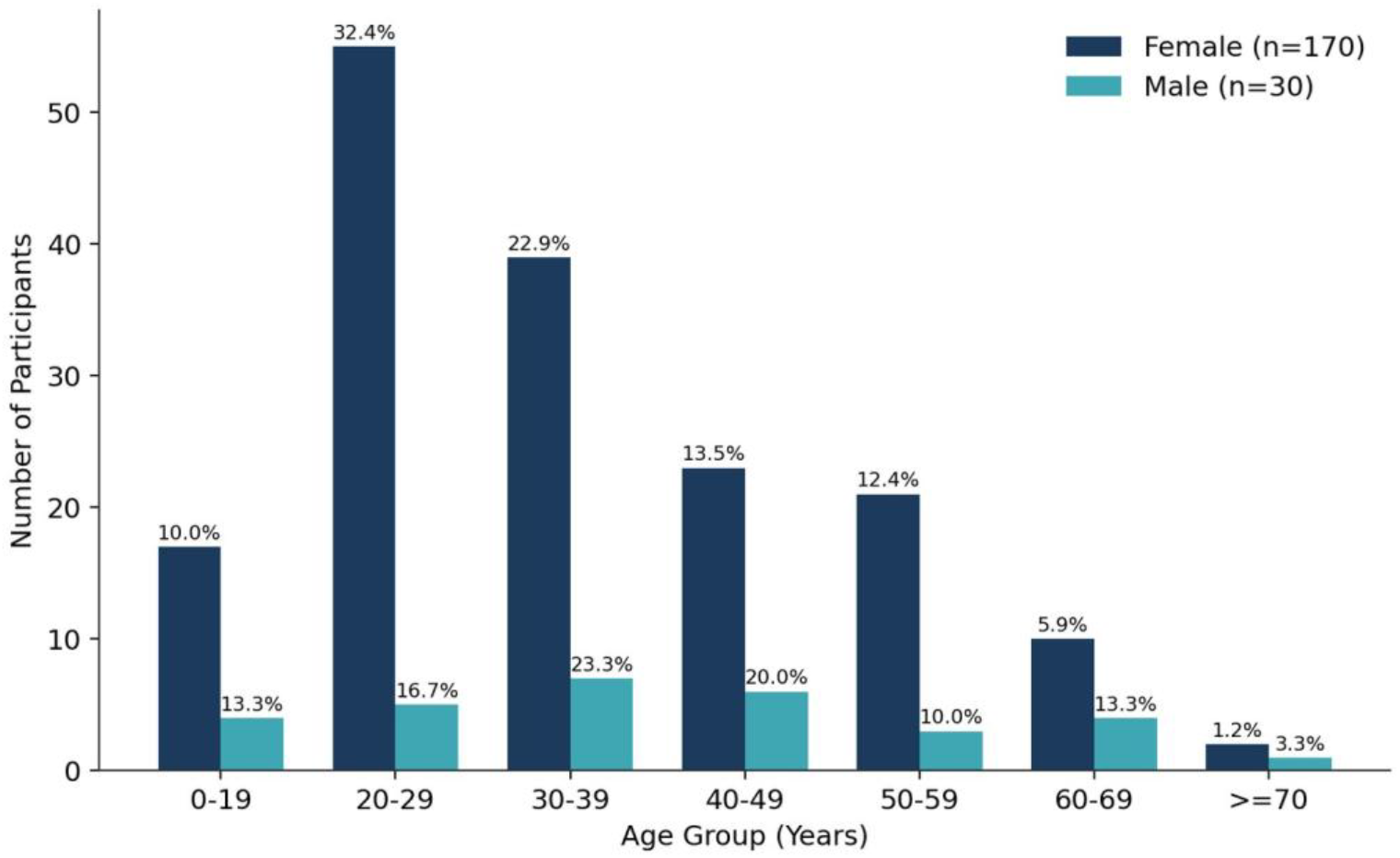
Age and sex distribution of study participants (N=200) among hospital outpatients in Jashore District, Bangladesh, October–December 2017. Bars represent the number of participants within each sex across seven predefined age categories, with percentages calculated within each sex group shown above the bars.

**Figure 2.**
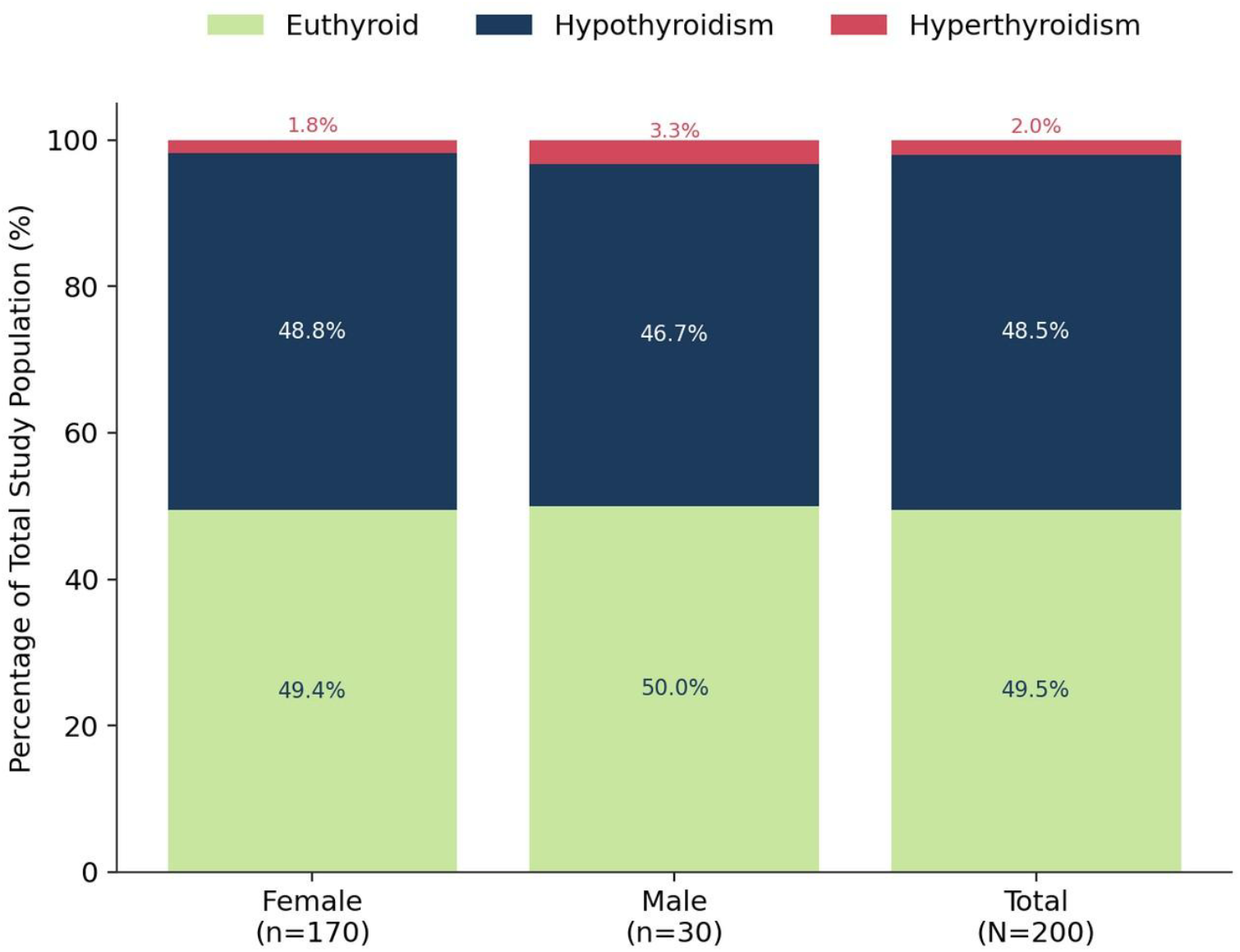
Thyroid status distribution by sex among hospital outpatients in Jashore District, Bangladesh, October– December 2017. Stacked bars depict the euthyroid, hypothyroid, and hyperthyroid proportions, expressed as a percentage of each group (female, male, total).

Among the participants (N=200), the 20–29-year group was the most numerically represented (31%, 62/200), followed by the 30-39-year group (24%, 47/200) (**Figure 1**). Age group showed a highly significant association with TSH abnormality (P<0.001, **Table 1**). Within-group prevalence of thyroid dysfunction peaked in the 40-49-year group (89.66%, 26/29), with the 50-59-year group ranking second (75.00%, 18/24), while the lowest within-group prevalence was observed in the 20-29-year group (29.03%, 18/62) (**Figure 3, Table 1**).

**Figure 3.**
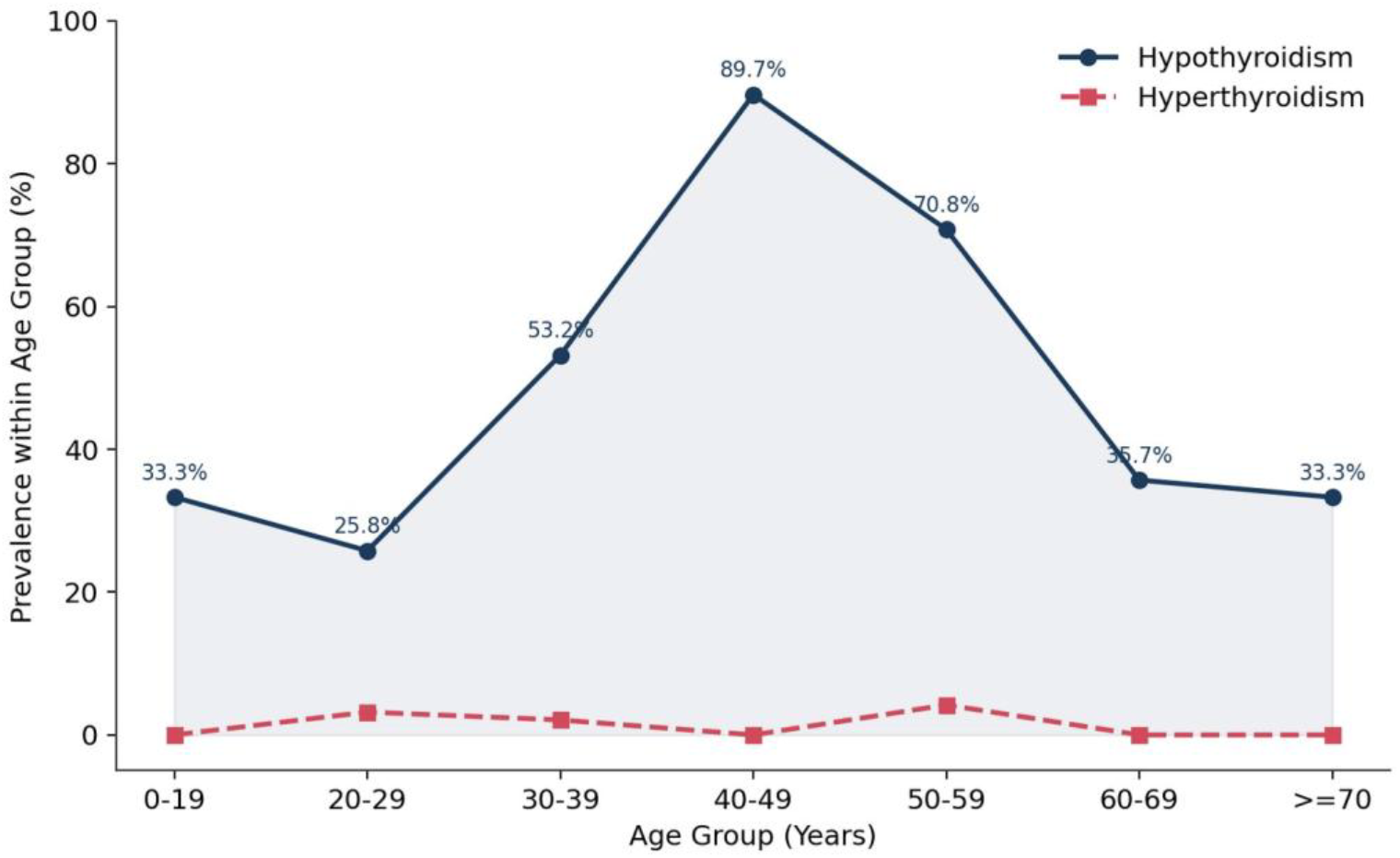
Age-stratified prevalence of hypothyroidism and hyperthyroidism among hospital outpatients in Jashore District, Bangladesh, October–December 2017. The y-axis represents the within-group prevalence (%) and the x-axis represents individual age groups (years).

In terms of absolute case burden rather than within-group rate, young adult females aged 20-29 years represented the largest single subgroup of hypothyroid cases, given their high representation in the overall sample. Childhood cases (0-19 years) were identified in both sexes, with an overall within-group prevalence of 33.3% (7/21), underscoring the occurrence of early-onset thyroid dysfunction. Hyperthyroidism prevalence was low throughout and did not show a consistent age trend, remaining under 5% in every age group (**Figure 3**).

Individually, hypothyroidism occurred more frequently than hyperthyroidism across all age and sex strata, confirming that the hypothyroid state was the predominant thyroid-health pattern in this population (**Table 2, Figure 2**).

## Discussion

According to our study, 50.5% of the total outpatient population showed an abnormal TSH value, which is notably elevated relative to community-based estimates that typically report TSH abnormality rates of 10-20% in South Asian populations (5, 9). This discrepancy most likely reflects selection bias inherent to a clinic-based design, in which participants had self-presented for thyroid evaluation, enriching the sample for symptomatic individuals. This is an important contextual limitation when extrapolating findings to the general population; nevertheless, the relative distribution of disorder types and demographic patterns provides clinically meaningful insight.

Females comprised the large majority of our cohort (85%) and carried the majority of both hypothyroid and hyperthyroid cases in absolute terms, consistent with well-established evidence that females are four to ten times more susceptible to thyroid disorders than males, a disparity attributed to estrogen-mediated potentiation of thyroid autoimmunity and the additive immunological challenges of reproductive events including menarche, pregnancy, and menopause (6, 14, 15). Notably, when we compared the within-sex prevalence directly, the rate of thyroid dysfunction was almost identical between females (50.6%) and males (50.0%), and this difference was not statistically significant (P=1.00). This suggests that, within this clinic-referred population, female predominance in the disease burden was driven largely by the much greater number of female patients presenting for evaluation rather than an intrinsically higher biological risk in this particular sample. This should be interpreted cautiously alongside the broader literature on sex-based susceptibility.

The concentration of hypothyroid cases among young adult females (20-39 years) is of particular clinical significance. Undiagnosed hypothyroidism in women of reproductive age is associated with menstrual irregularity, infertility, and adverse pregnancy outcomes, including impaired fetal neurodevelopment (16). These findings underscore the need for routine thyroid screening among reproductive-age women attending health facilities in Bangladesh.

The identification of thyroid dysfunction in the 0-19-year age group (33.3% within-group prevalence) highlights the clinical importance of pediatric thyroid surveillance. Congenital hypothyroidism and juvenile autoimmune thyroiditis are the leading etiologies of thyroid dysfunction in children and adolescents, and undetected hypothyroidism during critical neurodevelopmental windows can result in growth retardation, psychomotor delay, and irreversible intellectual impairment (17).

The marked rise in within-group prevalence through middle adulthood, peaking at 40-49 years (89.7%) and remaining high at 50-59 years (75.0%), and the highly significant association between age group and TSH abnormality (P<0.001), suggest that middle-aged adults constitute a high-risk group warranting targeted screening. Several factors may contribute to this pattern, including the increasing prevalence of metabolic comorbidities mechanistically linked to thyroid dysfunction, as well as age-related changes in thyroid gland architecture and reduced thyroid reserve (8, 18).

The evolution of thyroid disease etiology in Bangladesh from iodine deficiency to autoimmune thyroid disease mirrors epidemiological transitions observed in other South Asian countries following iodine supplementation programs (10, 20). This transition necessitates a corresponding shift in diagnostic and management strategies from goiter prevalence surveys toward systematic TSH-based and antibody-based screening programs.

Like all studies, our study has some limitations. The cross-sectional design precludes causal or temporal inference. Clinic-based recruitment introduces selection bias that limits generalizability to the broader community. TSH measurement alone, without concurrent free T3, free T4, or thyroid autoantibody assessment, precludes etiological classification of thyroid dysfunction. The relatively small sample size (n=200) from a single center limits statistical power for finer subgroup analyses, and self-reported medical histories are susceptible to recall bias. However, this pilot study gives an idea of the prevalence of TSH abnormalities in Jashore City which will be applicable towards public health concern.

## Conclusion

In conclusion, the overall prevalence of TSH abnormalities among hospital outpatients in Jashore district, Bangladesh, is quite high (about 50.5%), driven principally by hypothyroidism, with prevalence rising sharply through middle adulthood and peaking at 40-49 years. Our findings provide baseline data on TSH abnormalities within the Jashore area that may inform targeted screening strategies for reproductive-age women and middle-aged adults. Additional studies, including community-based and larger, multi-center surveillance with comprehensive thyroid biochemical panels, are necessary to understand the comprehensive public health burden of thyroid dysfunction in Bangladesh.

## Data Availability

All data produced in the present study are available upon reasonable request to the authors

## Acknowledgment

We are grateful to the clinical and laboratory staff of LabAid Hospital, Jashore Branch, Bangladesh, for their cooperation throughout data collection. We thank all study participants for their voluntary contribution and the Department of Microbiology at Jashore University of Science and Technology (JUST) for institutional support.

## Ethics of Study

The study received ethical approval from the Ethical Review Committee of Jashore University of Science and Technology (JUST), Jashore, Bangladesh. Written informed consent was obtained from all adult participants; for participants under 18 years of age, parental or guardian consent was secured alongside participant assent, in accordance with the Declaration of Helsinki.

## Funds

This research received no specific grant from any funding agency in the public, commercial, or not-for-profit sectors.

